# Observational Study on 255 Mechanically Ventilated Covid Patients at the Beginning of the USA Pandemic

**DOI:** 10.1101/2021.05.28.21258012

**Authors:** Leon G. Smith, Nicolas Mendoza, David Dobesh, Stephen M. Smith

## Abstract

**Introduction:** This observational study looked at 255 COVID19 patients who required invasive mechanical ventilation (IMV) during the first two months of the US pandemic. Through comprehensive, longitudinal evaluation and new consideration of all the data, we were able to better describe and understand factors affecting outcome after intubation.

**Methods:** All vital signs, laboratory values, and medication administrations (time, date, dose, and route) were collected and organized. Further, each patient’s prior medical records, including PBM data and available ECG, were reviewed by a physician. These data were incorporated into time-series database for statistical analysis.

**Results:** By discharge or Day 90, 78.2% of the cohort expired. The most common pre-existing conditions were hypertension, (63.5%), diabetes (59.2%) and obesity (50.4%). Age correlated with death. Comorbidities and clinical status on presentation were not predictive of outcome. Admission markers of inflammation were universally elevated (>96%). The cohort’s weight range was nearly 7-fold. Causal modeling establishes that weight-adjusted HCQ and AZM therapy improves survival by over 100%. QTc prolongation did not correlate with cumulative HCQ dose or HCQ serum levels.

**Discussion:** This detailed approach gives us better understanding of risk factors, prognostic indicators, and outcomes of Covid patients needing IMV. Few variables were related to outcome. By considering more factors and using new methods, we found that when increased doses of co-administered HCQ and AZM were associated with >100% increase in survival. Comparison of absolute with weight-adjusted cumulative doses proves administration ≥80 mg/kg of HCQ with > 1 gm AZM increases survival in IMV-requiring Covid patients by over 100%. According to our data, HCQ is not associated with prolongation. Studies, which reported QTc prolongation secondary to HCQ, need to be re-evaluated more stringently and with controls.

The weight ranges of Covid patient cohorts are substantially greater than those of most antibiotic RCTs. Future clinical trials need to consider the weight variance of hospitalized Covid patients and need to study therapeutics more thoughtfully.

## Introduction

Several studies have reported the relationship of factors or variables, measured at admission, to mortality rate of hospitalized, Covid patients (1–8). However, fewer studies have focused on critically ill Covid patients (9). Fewer still focus on Covid patients, requiring invasive mechanical ventilation (IMV). In a retrospective, observational study, we examined the associations of comorbidities, laboratory parameters, and medical therapies with mortality in this critically ill population.

In northern New Jersey, the first wave of this pandemic began in the second week of March and peaked in early April 2020. This cohort consists of all Covid patients, who were admitted to a single hospital during this initial wave of the pandemic and who required intubation and invasive mechanical ventilation (IMV). All patients except one, who nosocomially acquired Covid, were admitted to Saint Barnabas Medical Center over a 51-day period from March 12^th^ through May 1^st^. Each datum from each patient’s medical record was extracted into a single database. Data were analyzed in various ways, including several not used before in observational Covid studies. The approaches included time series analysis of vital signs, laboratory values and therapeutics. We also examined therapeutics in a more thorough, comprehensive manner, which included all therapeutics used. In doing so, we were able to go new insights into severe Covid, causing respiratory failure.

## Methods

### Study population & Data Extraction

Saint Barnabas Medical Center is a 557-bed, teaching medical center in Livingston, New Jersey. Using the hospital’s discharge coding data, we identified 255 Covid patients, who were admitted by May 1, 2020 and required invasive mechanical ventilation (IMV). Ethical approval for the study was granted by the hospital’s Institutional Review Board.

Data were extracted and combined into a database. Each patient’s data were combined into a dataset containing relevant time, date, result, and dosing information. In addition, each chart was reviewed to extract past medical history, prior medical attention, and outpatient medication use.

### Clinical Definitions

ADA definitions were used to diagnose DM, PreDM and stress hyperglycemia (10,11). To evaluate corticosteroid therapy, each dose was converted to the equivalent dose of dexamethasone. Acute kidney injury (AKI) was defined according to selected criteria from the Kidney Disease: Improving Global Outcomes definition of Stage 3 AKI (12).

Each electrocardiogram was manually reviewed by an electrophysiologist. The corrected QT interval was calculated using the Bazett formula. In patients with a prolonged QRS, the calculated QTc was corrected by the degree to which QRS duration exceeded 120msec.

### Statistical Analyses

The Student t-test, Whitney-Mann U test, and Chi-squared test were used for comparison of means, medians, and proportions, respectively.

Descriptive statistics were used to report patients ‘ characteristics. Multiple logistic regressions were performed to assess what factors were associated with patients ‘ survival status. Odds ratio (OR) and 95% confidence interval (95% CI) were reported. Then, the survival curve was plotted through Kaplan-Meier on the medicine variables. To examine what factors impacted mortality risk over the 60 days of admitting in hospital, a series of cox proportional hazard regression analyses were conducted. Hazard ratio (HR) and 95% CI were then calculated. All data were analyzed using MedCalc, R software with survival, survminer, lme4, and ggplot2 packages. Statistical significance was expressed as a *p* < .05.

A causal model was developed to provide study of relationships between assigned variables related to the study of critically ill Covid patients that utilized the framework of Rubin’s Potential Outcome Model or the Rubin-Neyman Model (13). Thus, the model’s main outcome Average Treatment Effect estimate (ATE) was calculated over multiple definitions of treatment. The ATE measures the difference in statistical mean outcomes between units assigned to the treatment as well as units assigned to the control.

We analyzed the covariates unbalanced and applied corrective trimming to re-balance the different factors according to their respective propensity scores. Thus, the analysis was calculated with the ATE using stratification by dividing the data set into blocks that have propensity score equivalent and computing individual treatment estimates and averaging them.

## Results Section

### Demographics

Using discharge coding records of >1,200 total Covid patients admitted over this 8-week period, we identified 255 patients, who required IMV. Mirroring the pattern of total Covid admissions, the admission rate for this cohort had a broad peak from late March through mid-April 2020 (Figure 1). 252 patients (98.8%) tested positive for SARS-CoV-2 RT-PCR or antigen assay during the admission or the prior week; 3 patients (1.2%) were diagnosed by their physicians, based upon clinical and laboratory findings. 63.6% of the cohort was ≥ 60 years-old (Table 1). The mean duration of symptoms prior to admission was 6.7 days (range: 1-30 days). The pre-existing conditions, found in at least 3.5% of the Cohort, are listed in Table 1. The most common, known comorbidities were hypertension (63.5%), diabetes (45.5%), hyperlipidemia (31.0%) and coronary artery disease (13.3%). 7.5% of the cohort had ESRD.

**Table 1:** Demographics, Presentation Values & Pre-existing Conditions.

| Characteristic | All | Alive | Expired |
| --- | --- | --- | --- |
| No. Pts | 255 | 54 | 201 |
| **Age** |  |  |  |
| All - No. (Range) | 63.7 (25-94) | 53.6 (30-86) | 66.4 (25-94) |
| < 60 - No. (%) | 92 (36.1%) | 37 (68.5%) | 55 (27.4%) |
| ≥ 60 - No. (%) | 163 (63.6%) | 17 (31.5%) | 146 (72.6%) |
| **Gender - No. (%)** |  |  |  |
| Female | 107 (42.0%) | 19 (35.2%) | 88 (43.8%) |
| Male | 148 (58.0%) | 35 (64.8%) | 113 (56.2%) |
| **Race and Ethnicity-No. (%)** |  |  |  |
| Black | 126 (40.4%) | 29 (53.7%) | 97 (48.3%) |
| White | 50 (19.6%) | 7 (13.0%) | 43 (21.4%) |
| Hispanic | 31 (12.2%) | 9 (16.7%) | 22 (10.9%) |
| Other | 30 (11.8%) | 7 (13.0%) | 21 (10.4%) |
| Asian | 18 (7.1%) | 2 (3.7%) | 16 (8.0%) |
| **Pre-existing Conditions-No. (%)** |  |  |  |
| Hypertension | 162 (63.5%) | 28 (51.9%) | 134 (66.7%) |
| Hyperlipidemia | 79 (31.0%) | 8 (14.8%) | 71 (35.3%) |
| Coronary artery disease | 34 (13.3%) | 3 (5.6%) | 31 (15.4%) |
| Congestive heart failure | 23 (9.0%) | 1 (1.9%) | 22 (10.9%) |
| Asthma | 20 (7.8%) | 5 (9.3%) | 15 (7.5%) |
| End-Stage Renal Disease | 19 (7.5%) | 4 (7.4%) | 15 (7.5%) |
| Chronic Kidney Disease Stage III | 18 (7.1%) | 3 (5.6%) | 15 (7.5%) |
| Atrial fibrillation | 17 (6.7%) | 2 (3.7%) | 15 (7.5%) |
| COPD | 15 (5.9%) | 0 (0%) | 15 (7.5%) |
| Cerebrovascular accident | 13 (5.1%) | 0 (0%) | 13 (6.5%) |
| Dementia | 12 (4.7%) | 0 (0%) | 12 (6.0%) |
| Transplant, Renal | 12 (4.7%) | 2 (3.7%) | 10 (5.0%) |
| Active Cancer | 9 (3.5%) | 0 (0%) | 9 (4.5%) |
| **Presentation Values** |  |  |  |
| BMI - Ave (Range) | 31.4 (16.3-65.7) | 32.2 (19.7-51.5) | 31.2 (16.3-65.7) |
| HbA1C - Ave % (Range) | 7.7 (4.1-16.4) | 7.7 (4.8-13.6) | 7.7 (4.1-16.4) |
| Glucose - Ave mg/dL (Range) | 199.3 (62-1,226) | 188.2 (68-1,129) | 202.2 (62-1,226) |
| T ≥100°F - No. (%) | 75 (29.4%) | 16 (29.6%) | 53 (26.4%) |
| Hypoxia on room air - No. (%) | 220 (86.3%) | 49 (90.7%) | 171 (85.1%) |

**Fig 1.**
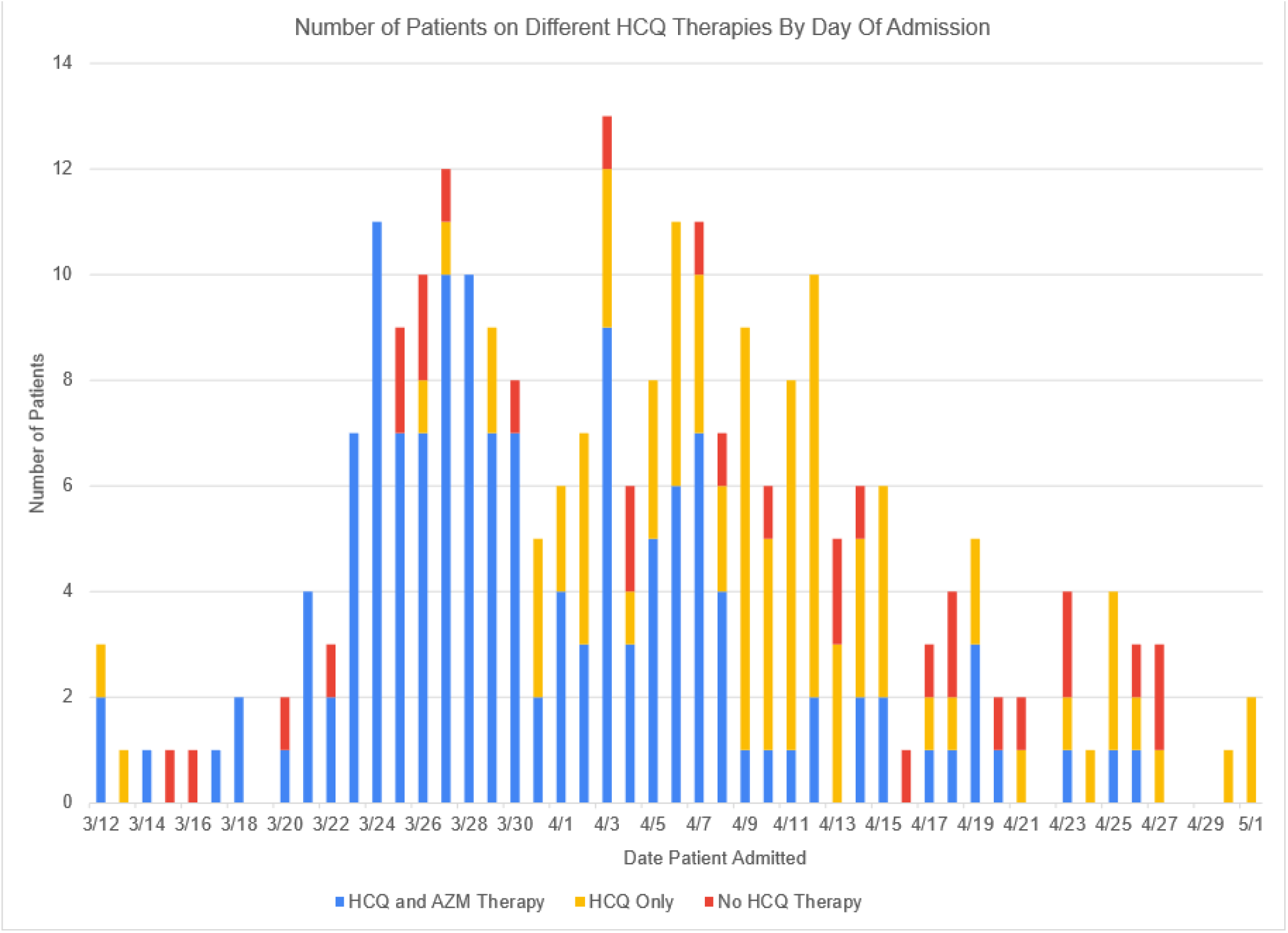
Number of patients by Date of Admission and breakdown by treatment with HCQ/AZM, HCQ alone or no HCQ therapy. Shown are the number of patients in the Cohort by admission date, from March 12 – May 1, 2020. HCQ therapy for each patient is demonstrated by use of color. Blue means the patient received HCQ and AZM therapy together, Gold, HCQ therapy without AZM, and Red, the patient did not receive HCQ.

Patients were observed to hospital discharge or Day 90 of hospitalization. 201 patients (78.8%) died prior to the end of observation (“Expired group”). 54 patients (21.1%) survived (“Alive group”). The median and mean lengths of stay = 13.1 and 19.2 days respectively. 53 patients (23.1%) had a LOS > 28 days. Of note, 7 patients (13.0%) of the Alive group were transferred prior to a long-term acute care hospital still on a ventilator and critically ill. 6 of these 7 patients were transferred to LTACH between Day 39-67. However, 1 pt was transferred on Day 18 of hospitalization. The Day 90 outcomes of these 7 patients are not known. Consequently, the mortality rate may be greater.

Surprisingly, few parameters distinguished the Alive and Expired groups. However, all 43 patients (16.7%) with active cancer, dementia, s/p CVA, and/or COPD expired. However, other comorbidities, including HTN, CAD, and ESRD, were not associated with worse outcome. Sex, race, clinical status on presentation and blood type also did not correlate with survival. For instance, the Alive group’s mean NEWS2 score (8.61) was slightly higher than the Expired group’s NEWS2 mean (8.52). Age and HLD correlated with outcome.

40.2% sought prior medical attention (PMA) ≥1 day prior to being admitted. PMA patients had similar NEWS2 scores (8.5 v. 8.6), a longer duration of symptoms in PMA patients was greater (7.5 v. 6.1 days) and lower survival rate (17.6% v. 23.5%) than patients admitted upon initial presentation.

### Laboratory Values

Ferritin, D-dimer, LDH and C-reactive protein (CRP) were measured in >76% of the Cohort near or at admission (Table 2.a). Over 96% of each parameter was greater than the ULN. The initial values of D-dimer, ferritin, and CRP in the Alive and Expired groups were similar. However, all 3 patients with an initial D-dimer >69,000 ng/ml expired. The mean LDH values was lower in the Alive than Expired group = 449 and 702 U/L respectively [p = 0.021].

**Table 2.** Admission Lab Values (a) and Slopes of Lab Values during Hospitalization (b).

|  | **Ferritin** | **D-dimer** | **LDH** | **CRP** |
| --- | --- | --- | --- | --- |
| No. Tested | 221 | 195 | 192 | 219 |
| Percent of values > ULN* | 98.2 | 96.5 | 99.0 | 99.5 |
| Ave value* | 2,701 | 8,968 | 640 | 16.2 |
| Maximum value* | 77,015 | >69,000 | 8,130 | 53.6 |

\* - The ULN (upper limit of normal) and unit of expression for ferritin = 150 ng/ml, D-dimer = 243 ng/mL, LDH = 200 U/L and CRP = 0.5 mg/dL.

b)
| **Lab** | **All** | **Expired** | **Alive** | **Difference** |
| --- | --- | --- | --- | --- |
| Ferritin Slope* | 76.4 | 102.8 | 3.5 | 99.3 (96.6%) |
| D-dimer Slope* | 38.6 | 187.3 | -315 | 502.3 (262.2%) |
| LDH Slope* | 38.6 | 54.9 | -2.6 | 57.5 (104.4%) |

\* Slope, calculated as unit change per day, was considered as an intuitive path of inquiry for prognostic indicators. These three labs' increase correlated with Covid19 disease. In turn, their first derivative, with dt equal to 1 day, produced promising leads for prognostics.

We hypothesized that the patterns of these lab values over time correlated with outcome. To test this hypothesis, we calculated, the slopes for the ferritin, D-dimer, and LDH levels. For each parameter, the slope of the Expired group was significantly higher than the slope for the Alive group (Table 2.b).

Acute kidney injury (AKI) developed in 77.5% of patients. Hyponatremia was present in 52.5% upon admission. 10.2% with AKI received renal replacement therapy (RRT). AKI was more common in the Expired group (83.3%) than the Alive (56.0%) group [p < 0.001]. Hyponatremia on presentation, however, was not associated with outcome. This survival rate for AKI patients, who received RRT, was 20.8% and similar to the overall survival rate. However, 40% permanently required RRT.

### Time of Intubation and Days on Ventilator

Most (57.3%) of the patients were intubated within 72 hours of arrival to the hospital. The average time to intubation after arrival was 4.1 days. The mean times of intubation for the Alive group (3.4 days) and Expired group (4.3 days) were similar. Accordingly, survival rates did not differ significantly by day of intubation. However, only 1 of the 16 patients, who were intubated on ≥ Day 15, survived. The median for pt days on IMV = 8.0 days. 55 patients (21.6%) were on IMV for >= 20 days.

### Hyperglycemia, DM and Obesity

45.5% of the Cohort had a history of DM. The mean A1C of known diabetics = 8.7% (range: 4.1-16.4%). An A1C was measured on 46.8% (65 of 139 pts) with no history of DM. The majority (90.8%) had an elevated A1C diagnostic of DM (53.8%) or PreDM (36.9%). 9.2% had a normal A1C (Table 3).

**Table 3.**
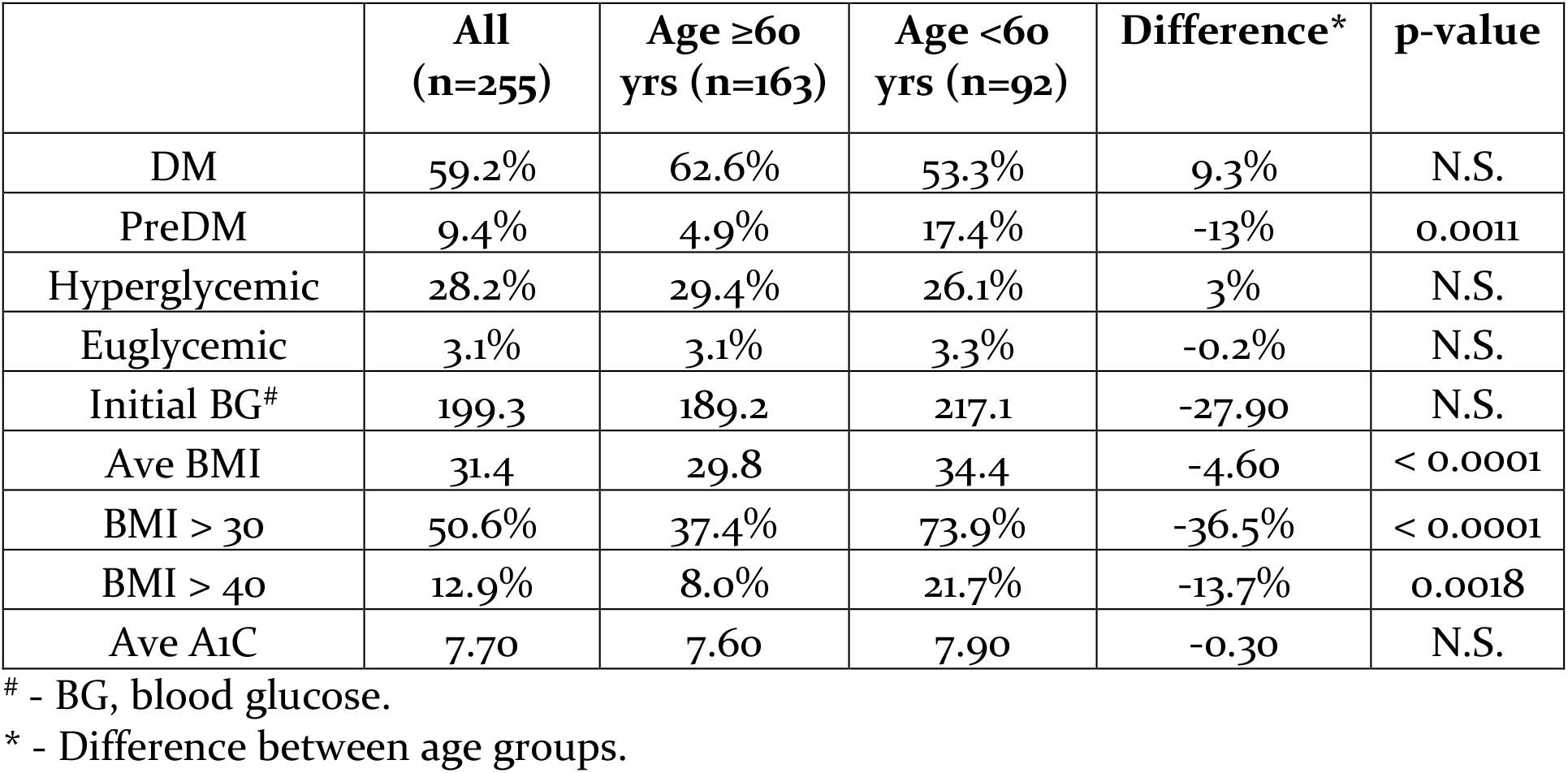
Diabetes Status, Obesity Rates, and HbA1C Levels by Age.

|  | All<br>(n=255) | Age ≥60<br>yrs (n=163) | Age <60<br>yrs (n=92) | Difference* | p-value |
| --- | --- | --- | --- | --- | --- |
| DM | 59.2% | 62.6% | 53.3% | 9.3% | N.S. |
| PreDM | 9.4% | 4.9% | 17.4% | -13% | 0.0011 |
| Hyperglycemic | 28.2% | 29.4% | 26.1% | 3% | N.S. |
| Euglycemic | 3.1% | 3.1% | 3.3% | -0.2% | N.S. |
| Initial BG<sup>#</sup> | 199.3 | 189.2 | 217.1 | -27.90 | N.S. |
| Ave BMI | 31.4 | 29.8 | 34.4 | -4.60 | < 0.0001 |
| BMI > 30 | 50.6% | 37.4% | 73.9% | -36.5% | < 0.0001 |
| BMI > 40 | 12.9% | 8.0% | 21.7% | -13.7% | 0.0018 |
| Ave A<sub>1</sub>C | 7.70 | 7.60 | 7.90 | -0.30 | N.S. |

The initial blood glucose (BG) was > 125, >140 and >200 mg/dL in 72.9%, 60.4% and 31.8%, respectively. 3 patients (1.2%) presented with a BG < 70 mg/dL. During the admission, DM and PreDM patients consistently had BGs ≥140 mg/dL, regardless of corticosteroid therapy.

The BG values of 80 patients with no history of DM and either a normal A1C (6 pts) or no A1C measurement (74 pts) were analyzed. For each, the maximal BG and the percent of BGs ≥140 mg/dL were determined. Corticosteroid therapy was noted. While off corticosteroids, 68 (72.5%) of 80 patients had a maximal BG >200 and BGs consistently >140 mg/dL. At some point during the admission, 63.6% of the 22 patients without significant hyperglycemia received corticosteroid therapy, resulting in hyperglycemia in 85.7%.

In total, only 10 patients (3.9%) of the cohort consistently had BGs <140 mg/dL. Conversely, the vast majority (96.1%) had high BG levels above 140 mg/dL and/or peak BG >200 mg/dL. Excluding corticosteroid-associated hyperglycemia reduces that percentage to 91.4%.

The DM status of 74 patients (29.0%) is not known. Based upon each pt’s BG analysis and the A1C results on patients with no history of DM, the majority of these 74 patients likely had impaired glucose metabolism prior to developing Covid.

The Cohort’s mean BMI = 31.4 (range: 16.3 – 65.7). 128 patients (50.4%) were obese, 81 (31.8%) were overweight and 46 patients (18.1%) had a normal BMI (Table 4). Patient < 60 years-old had a greater mean BMI (34.4) than patients ≥60 years-old (29.8) [p < 0.0001]. Accordingly, obesity was more common in the <60-year-old group (73.9% v. 37.4%; p < 0.0001).

**Table 4:** Cox Proportional Hazard Regression to Assess Factors Affecting the Survival Status.

| *Covariate* | *HR* | *SE* | *CI* | *t* | *p* |
| --- | --- | --- | --- | --- | --- |
| Age [Old]* | 1.92 | 0.19 | 1.32 - 2.77 | 3.46 | **0.0005** |
| AFIB<sup>a</sup> | 1.48 | 0.3 | 0.82 - 2.68 | 1.30 | 0.19 |
| Asthma<sup>a</sup> | 1.20 | 0.29 | 0.68 - 2.13 | 0.64 | 0.52 |
| CAD<sup>a</sup> | 0.99 | 0.24 | 0.62 - 1.58 | -0.04 | 0.97 |
| CHF<sup>a</sup> | 1.10 | 0.26 | 0.67 - 1.82 | 0.39 | 0.70 |
| CKD<sup>a</sup> | 0.82 | 0.29 | 0.46 - 1.45 | -0.68 | 0.49 |
| ESRD<sup>a</sup> | 1.24 | 0.16 | 0.91 - 1.69 | 1.34 | 0.18 |
| HLD<sup>a</sup> | 0.78 | 0.29 | 0.44 - 1.39 | -0.84 | 0.40 |
| HTN<sup>a</sup> | 1.14 | 0.19 | 0.78 - 1.66 | 0.67 | 0.50 |
| DM<sup>a</sup> | 1.14 | 0.17 | 0.82 - 1.59 | 0.77 | 0.44 |
| BMI | 1.00 | 0.01 | 0.97 - 1.02 | -0.33 | 0.74 |
| Sex [Male] | 0.88 | 0.16 | 0.63 - 1.21 | -0.81 | 0.42 |
| >3g HCQ/>1g AZM | 0.31 | 0.26 | 0.18 - 0.51 | -4.47 | **<0.0001** |
| Tocilizumab | 0.50 | 0.17 | 0.36 - 0.69 | -4.16 | **<0.0001** |
| Steroids<sup>#</sup> | 0.71 | 0.17 | 0.51 - 0.98 | -2.10 | **0.036** |

Note: Overall model $X^2(15) = 93.87$ , $p < 0.0001$ . N = 255. AFIB, atrial fibrillation; CAD, coronary artery disease; CHF, congestive heart failure; CKD, chronic kidney disease; ESRD, end-stage renal disease; HLD, hyperlipidemia; HTN, hypertension; DM, diabetes mellitus; BMI, body mass index; AZM, azithromycin; HCQ, hydroxychloroquine; Steroid, glucocorticoid exposure; HR, hazard ratio; SE, standard error; CI, confidence interval.
<sup>a</sup> - compared to “No”.
\* - Old ( $\geq 60$ years) versus Young ( $< 60$ years).
<sup>#</sup> - Steroid therapy defined as dexamethasone $\geq 6$ mg or equivalent dose for $\geq 3$ days.

The average weight of this cohort = 89.7 kg (S.D. = 25.2 kg; range = 30.8 – 210 kg). This greatest weight was 6.8 times the lowest. 41 patients (16.1%) weighed ≥2 times the weight of 20 patients (7.8%). There were no significant differences in outcome by diabetes or obesity class. The survival rates for DM, PreDM and nonDM/unknown DM patients were 19.2%, 29.2% and 22.5% respectively. The mean admission BG level for Alive patients was non-significantly lower than that of the Expired patients. The BMI and A1C means of the Alive and Expired groups were nearly identical (Table 1).

### Therapeutics

We performed extensive analyses of all therapeutics given to Cohort patients during these admissions. Initially, we studied therapeutics by drug class, such as benzodiazepine or calcium channel blocker. When a drug class showed a potential effect on outcome, we analyzed the individual drugs. Therapeutics were analyzed categorically, in time-series, fashion and/or by cumulative dose. Other than the medications discussed below, no medication or class of medication was associated with outcome.

### Corticosteroids

Each dose of hydrocortisone, prednisone, and methylprednisolone was converted to the equivalent dose of dexamethasone. A patient was then deemed as having received “steroids” on a given day if the dose or its equivalent was ≥6 mg dexamethasone (based on the dose used in the Recovery Trial).

133 patients (52.2%) received at least one day of ≥6 mg dexamethasone. Steroid therapy was assessed in several ways. We analyzed steroid therapy by days on therapy. Patients were divided into two based on days of steroid therapy and outcomes were evaluated in binary fashion. We compared ≥ 1 days to 0 days, ≥3 days to ≤2 days, and ≥10 days to ≤9 days. The results were inconclusive as discussed below.

The effects of steroids before and after intubation were analyzed as well. Specifically, 37 and 46 patients received at least 3 days of steroid therapy before or after intubation. Survival rates were similar. 50.2% of the Expired group and 37.0% of the Alive group received ≥3 days of steroid therapy.

### Tocilizumab

91 patients (35.7%) of the Cohort received 1-2 doses of tocilizumab (TOZ) and 35.2% survived. The mean age of patients who received TOZ was lower than that of those who did not [57.1 .v. 67.2 yrs; p < 0.0001]. The mean and median hospital days of TOZ administration were 6.1 and 5.3 days (range = Day 1-27).

### Convalescent plasma

Convalescent plasma (CP) became available during week 4 and was given to 50 patients (19.6%) of the Cohort. CP-receiving patients were younger than non-CP receiving patients [59.9 v. 64.6 yrs; p = 0.032]. The mean and median times of CP administration were 13.2 and 10.8 days. The survival rates of CP-receiving patients (34.0%) was greater than those who did not (18.1%).

### Hydroxychloroquine

Hydroxychloroquine (HCQ) (≥ 400 mg) was given to 224 patients (87.8%). The average age of patients, who received HCQ, was similar to those who did not [63.5 v. 64.6 yrs; p=0.69]. 86.1% and 94.2% received the first dose of HCQ within 24 and 48 hours of ER arrival, respectively. The majority (56.3%) received a cumulative HCQ dose = 2,000 – 3,000 mg and 62.5% of HCQ patients were also given AZM (Bar graph). The coadministration of AZM with HCQ declined during this 6-week period (Figure 1).

Initial lasso and Cox proportional hazard model regression analyses showed that higher cumulative doses of HCQ were associated with a lower mortality rate. With every natural log increase in HCQ cumulative dose, patients were 1.12 times less likely to die [p<0.001]. Accordingly, 3,000 mg HCQ cumulative dose had a survival OR = 2.46.

When AZM and HCQ were given together, the association with survival greater than when HCQ was given alone. We finally noticed that patients, who received cumulative doses HCQ > 3,000 mg and AZM > 1,000 mg, had a much higher survival rate than all others.

37 patients received > 3g HCQ and > 1g AZM. 18 patients (48.6%) of these 37 patients survived. Comparatively, 36 patients (16.5%) of 218 patients who received either <= 3g HCQ or <= 1g AZM survived. The absolute difference (32.1%) in survival was significant [C.I = 15.9% −48.2%; p <0.0001]. The relative difference in survival = 194.5%. Differences of these magnitudes have not been reported in other clinical studies.

To understand the contributions of all variables to outcome, multiple Cox and logistic regression analyses were run. CP had no demonstrable effect on survival, presumably because it was given late and only to a small percentage of the Cohort. An individual Survival Curve was plotted for HCQ/AZM, TOZ and steroid therapy. As shown in Figure 2, the plots demonstrate that the risk of mortality was different in whether or not a medicine was provided (>3gHCQ/>1gAZM, tocilizumab, and steroid).

**Figure 2.**
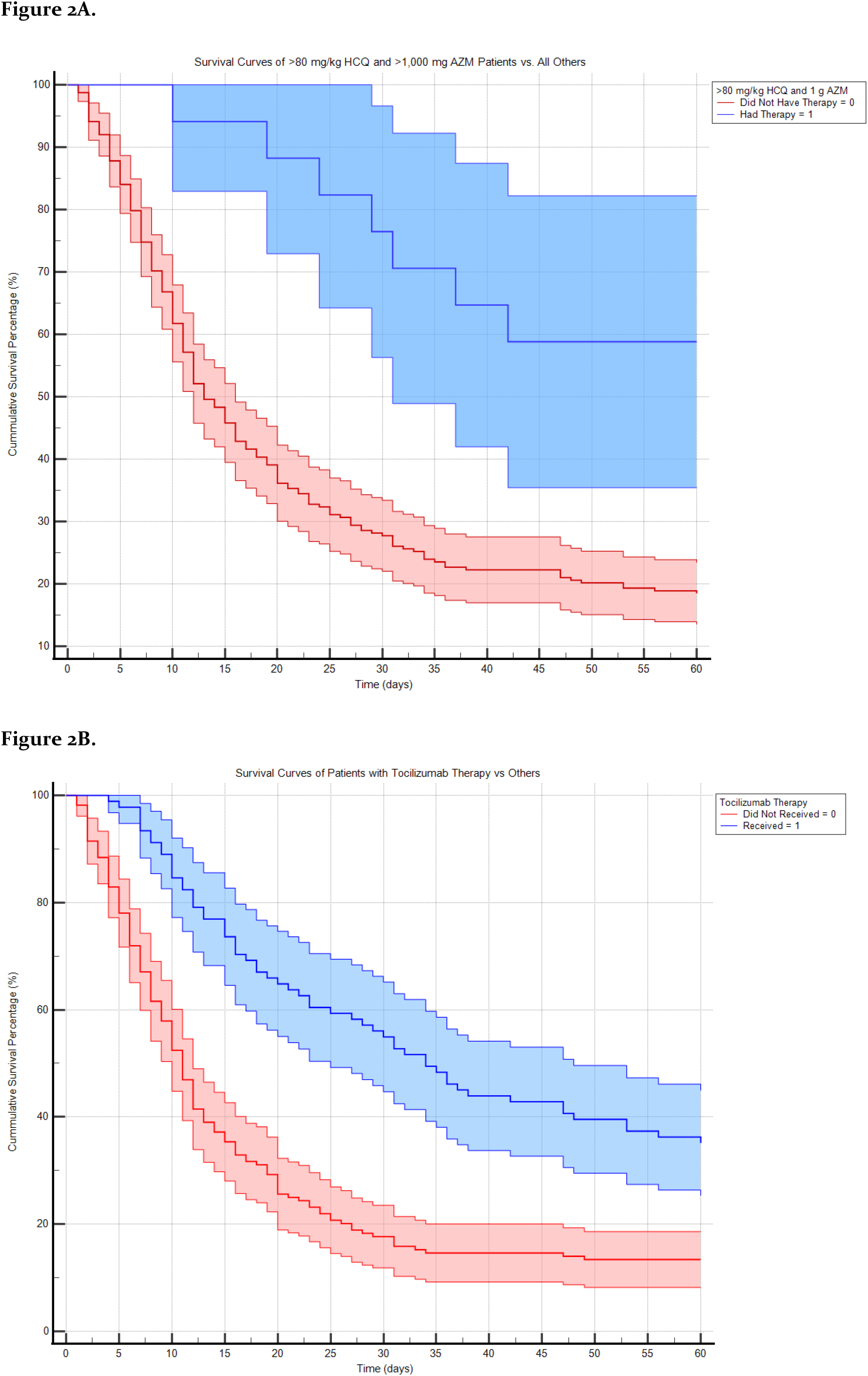
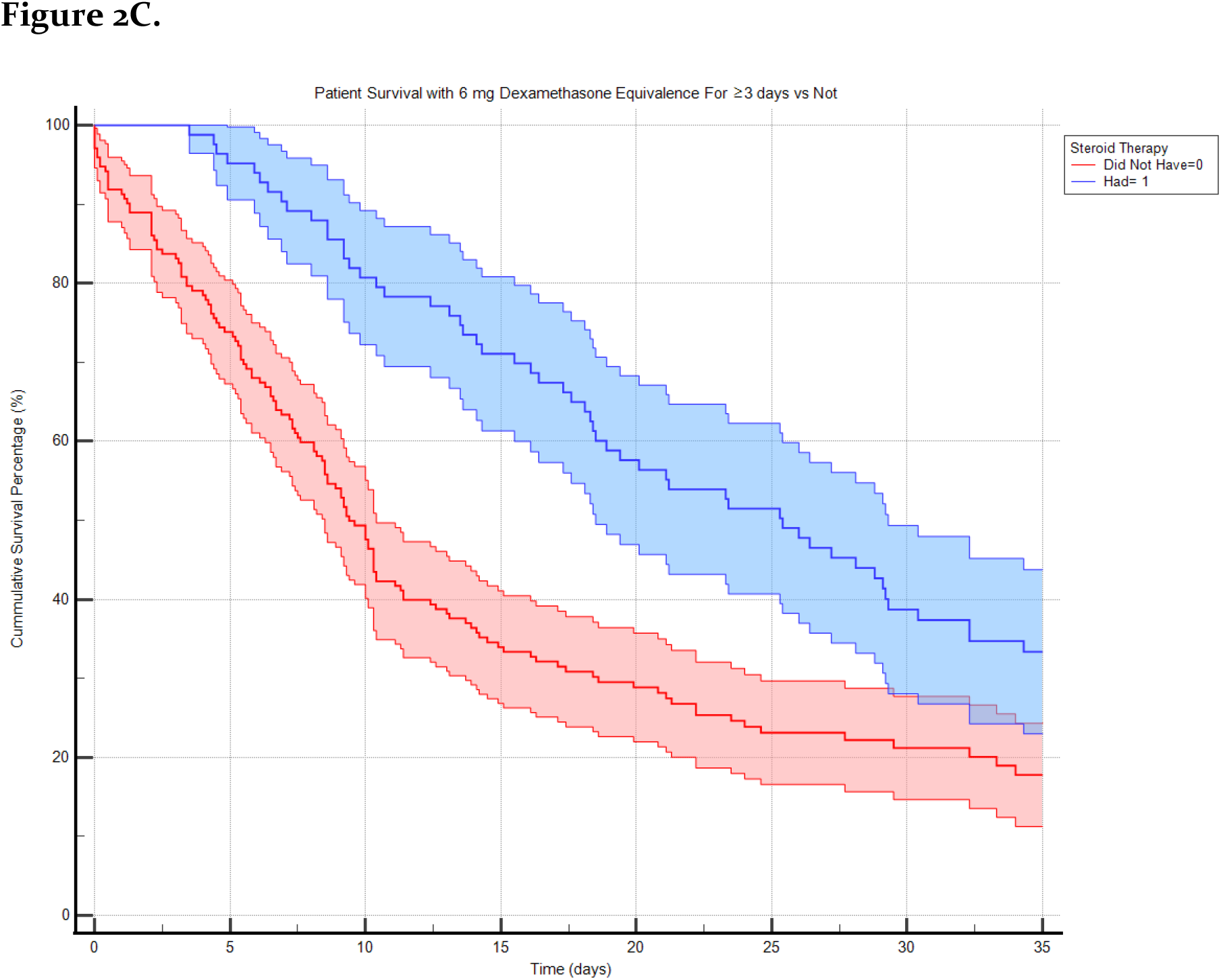
Cumulative survival curves with are shown for weight-adjusted, cumulative HCQ/AZM therapy (a), tocilizumab therapy, and steroid therapy (c). Blue line indicates receipt of therapy; Red line, no therapy. (95% CI shaded)

Then, survival analyses using multivariable cox proportional hazard regression were conducted to examine the mortality risk over 60 days in hospital when controlling for other covariates. The results (Table 4) show that the overall model was statistically significant, X2(15) = 93.87, p < .001.

Patients receiving >3g HCQ/>1g AZM had 3.26 times lower risk of mortality than those who did not receive these doses [HR = .307,95% CI = 0.183 – 0.516, p < .001]. Patients receiving TOZ had 2.00 times lower risk of death than patients who did not receive the medicine [HR = .50, 95% CI = 0.36 – 0.69, p < .001]. Additionally, patients, who received 6 mg or above steroid for at least 3 days, had 1.42 times lower risk of mortality [HR = .706, 95% CI = 0.510-0.978, p < .036].

To better understand the effect of HCQ/AZM therapy and outcome, we analyzed the data by Rubin’s Causal model. The Rubin model’s main outcome is an Average Treatment Effect (ATE) estimate. The ATE is a measure used to compare treatments (or interventions) in randomized experiments, evaluation of policy interventions, and observational medical trials. The ATE measures the difference in mean outcomes between units assigned to the treatment and units assigned to the control.

“Treatment” was defined as receipt of >3 gm HCQ and >1 gm AZM. 37 patients received the “Treatment”; 218 patients did not. The analysis revealed that the “Treatment” increases the absolute chances of survival by 22.5% [95% C.I. = 7.6 % – 37.5%; p = 0.003]. In other words, for the untreated group, survival was 16.5%. Treatment of this group with >3 gm HCQ and >1 gm AZM would have resulted in an increase in survival rate to 39.0% [95% C.I. 24.1% −44.0%, p = 0.003] or more than a 100% increase in survival rate. We then ran Rubin’s causal model, substituting other variables, such as blood type, in as the “Treatment” group. Blood Type was selected due to naï ve high correlation.

We found no statistically significant correlation with any other variable tested. To further explore the relationship between outcome and HCQ, we used similar analysis but considered HCQ cumulative dose as a variable. The range of the cumulative dosage is 0 – 8,000 mg. The line in Figure 3a shows ATE vs. cumulative HCQ dose. The shaded purple area indicates the corresponding 95% C.I. The ATE significantly abruptly increases at a cumulative dose = 3,000 mg and then reaches a maximal ATE (26.9%) at 5,600 mg cumulative HCQ dose.

**Fig 3.**
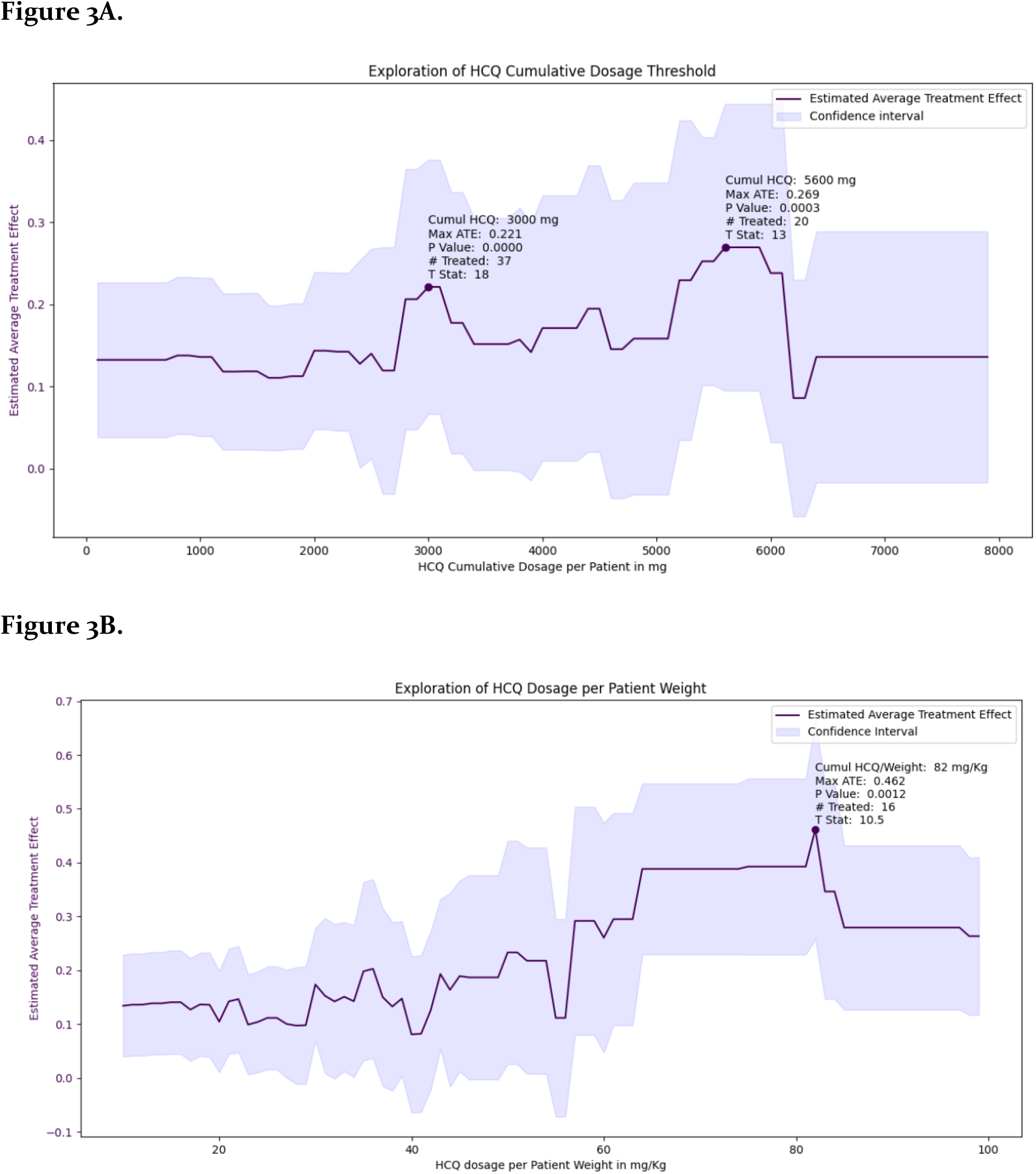
Average Treatment Effect of HCQ on survival Covid patients requiring IMV. ATE vs. cumulative HCQ dose and B). ATE vs. weight-adjusted HCQ cumulative dose. The Average Treatment Effect (ATE) increases with HCQ cumulative dose (A) peaking at 5,600 mg with a maximal ATE = 26.9%. Using weight-adjusted HCQ cumulative dose (B), the ATE peaks at 82 mg/kg of HCQ with a maximal ATE = 46.2%. (95% C.I. is shaded.)

We reasoned: if cumulative HCQ dose is causally linked to an increased chance of survival, then the weight-adjusted cumulative HCQ dose would have an even stronger effect on survival. The maximal weight in this cohort is 6.8 x the minimal weight. As shown in Figure 3b, higher cumulative, weight-adjusted HCQ doses produce greater ATE. The ATE increases suddenly between 40-50 mg/kg and reaches a maximal ATE = 46.2% at 82 mg/kg [p=0.0012].

For confirmation purposes, we performed logistic regressions on HCQ/AZM, TOZ and steroids. Further, we ram the regressions with two different definitions of HCQ therapy (AZM therapy was unchanged at >1 g). Based upon the ATE findings, HCQ therapy was defined as ≥80 mg/kg HCQ for the first analysis. Logistic regression demonstrates that the overall model was significant, X2(15) = 65.00, p < .001. The results (Table 5) reveal that patients receiving ≥80mg/kg HCQ and > 1g AZM were 14.18 times more likely to survive than those who did not [OR = 14.18, 95% CI = 4.05 – 55.61, p < .001].

**Table 5.** Factors Affecting the Survival Status of Patients with COVID-19 by Logistic Regression.

| Predictors | Survival |  |  |  |  |
| --- | --- | --- | --- | --- | --- |
|  | OR | SE | CI | Statistic | p |
| Age [Old]* | 0.2 | 0.08 | 0.08 – 0.43 | -3.93 | **0.0001** |
| Afib<sup>a</sup> | 0.65 | 0.65 | 0.07 – 3.92 | -0.43 | 0.671 |
| Asthma<sup>a</sup> | 2.12 | 1.35 | 0.57 – 7.27 | 1.18 | 0.238 |
| CAD<sup>a</sup> | 1.6 | 1.37 | 0.26 – 8.27 | 0.55 | 0.584 |
| CHF<sup>a</sup> | 0.52 | 0.59 | 0.03 – 3.48 | -0.57 | 0.566 |
| CKD<sup>a</sup> | 1.87 | 1.43 | 0.36 – 7.81 | 0.82 | 0.414 |
| ESRD<sup>a</sup> | 1.12 | 0.87 | 0.22 – 4.87 | 0.15 | 0.883 |
| HLD<sup>a</sup> | 0.29 | 0.17 | 0.08 – 0.87 | -2.05 | **0.040** |
| HTN<sup>a</sup> | 1.09 | 0.44 | 0.50 – 2.45 | 0.22 | 0.822 |
| DM<sup>a</sup> | 0.82 | 0.32 | 0.38 – 1.76 | -0.52 | 0.605 |
| BMI | 1 | 0.03 | 0.94 – 1.05 | -0.14 | 0.891 |
| Sex [Male] | 1.89 | 0.75 | 0.87 – 4.22 | 1.59 | 0.112 |
| $\geq 80$ mg/kg HCQ/ $> 1$g AZM<sup>a</sup> | 14.18 | 9.36 | 4.05 – 55.61 | 4.02 | **0.0001** |
| Tocilizumab<sup>a</sup> | 2.82 | 1.08 | 1.34 – 6.06 | 2.71 | **0.0067** |
| Steroids<sup>a#</sup> | 0.74 | 0.3 | 0.33 – 1.63 | -0.74 | 0.461 |

Note: Overall model $X^2(15) = 65.00$ , $p < 0.0001$ . Adjusted $R^2 = .256$ . $p < 0.0001$ . $N = 255$ .
AFIB, atrial fibrillation; CAD, coronary artery disease; CHF, congestive heart failure; CKD, chronic kidney disease; ESRD, end-stage renal disease; HLD, hyperlipidemia; HTN, hypertension; DM, diabetes mellitus; BMI, body mass index; AZM, azithromycin; HCQ, hydroxychloroquine; Steroid, glucocorticoid exposure, HR, hazard ratio; SE, standard error; CI, confidence interval.
<sup>a</sup> - compared to “No”.
\* - Old ( $\geq 60$ years) versus Young ( $< 60$ years).

Similarly, patients receiving tocilizumab were 2.82 times more likely to survive than those who did not receive tocilizumab [OR = 2.82, 95% CI 1.34 – 6.06, p = .007]. Furthermore, older patients (≥60 years old) were 5 times less likely to survive as compared to the young patients (<60 years old) [OR = .20, 95% CI 0.08 – 0.43, p < .001]. Patients with HLD were also 3.45 times less likely to survive than those who did not have HLD [OR = .29, 95% CI = 0.08 – 0.87, p = .040]. However, steroid therapy, defined as ≥6 mg dexamethasone for ≥3 days, was not significantly related to the survival status, p = .461. No other comorbidity or variable was associated with survival status.

While keeping everything else the same, we ran logistic regression analysis again with HCQ therapy defined as >3 g. Again, the overall model was significant, X2(15) = 65.78, p < .001. The odds ratio, 95% CI’s and p-values for the other variables were extremely similar to those from those in Table 5. However, the OR for >3 g HCQ = 7.03 or less than ½ the OR for ≥80mg/kg HCQ.

### Clinical Condition at Discharge

83.3% of the Alive group had not returned to pre-admission health status at discharge. 8 patients (14.8%) were still on IMV. 44.4% patients were discharged to home. 35.2% were unable to perform ADLs. 18 patients 33.3% had difficulty walking and/or required oxygen therapy. Only 16.7% of Alive group or 3.5% of the Cohort, were discharged to home, off oxygen therapy and without cognitive or motor deficit.

### Effects of HCQ +/-AZM on QTc

24 (47.1%) of the 51 patients whose total cumulative HCQ dose >3000 mg had ≥ 2ECGs performed while on HCQ. A total of 145 ECGs were interpreted by an electrophysiology cardiologist. The mean number of ECGs per patient on HCQ therapy = 6 with a maximum = 11.

For each ECG, the change (delta) in the QTc from baseline was calculated. The mean QTc delta = 16.9 ms (range = −79 −128 ms). As shown in Figure 4a, there was no correlation between HCQ cumulative dose and QTc delta (r = −0.170).

**Figure 4.**
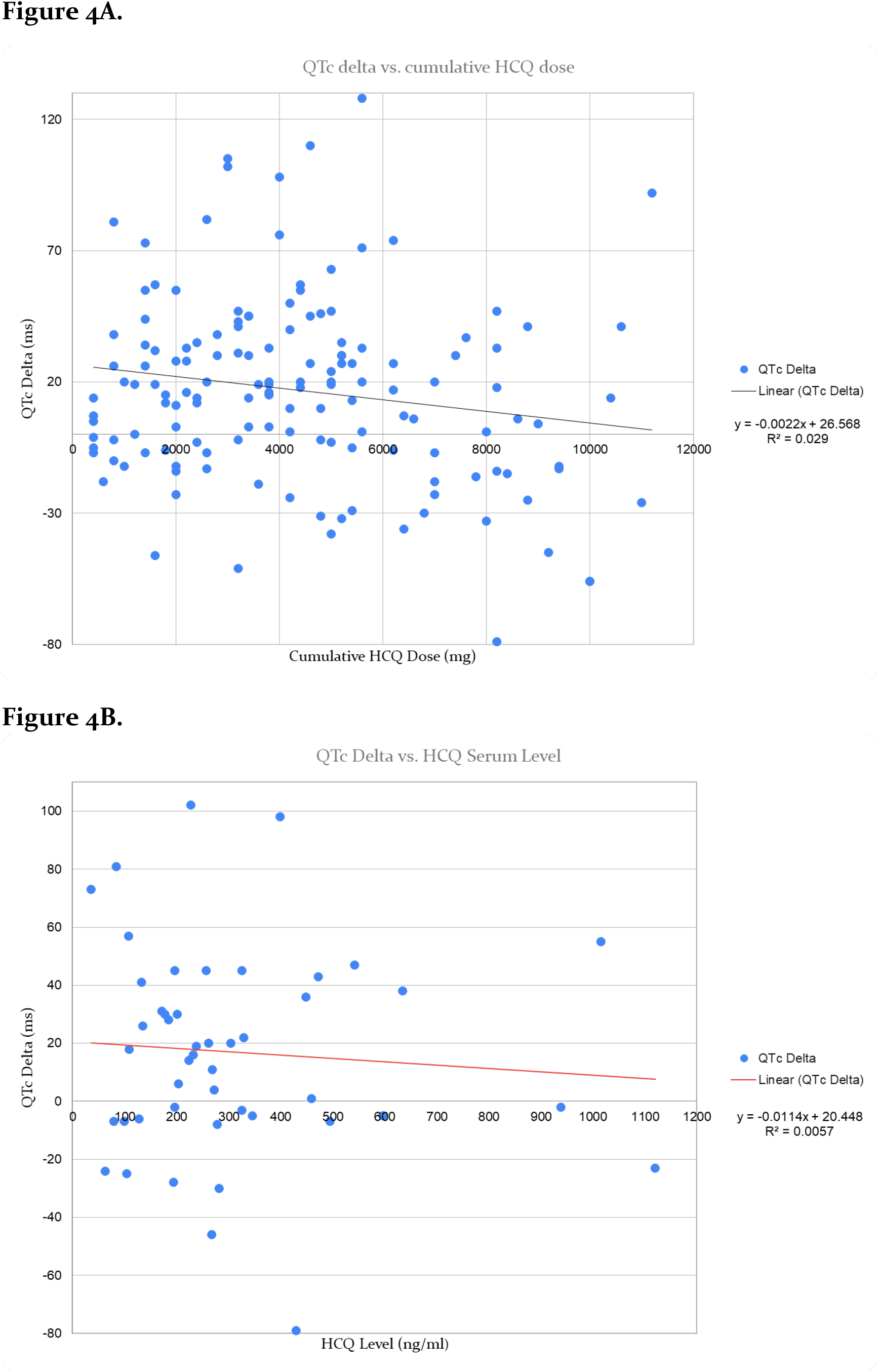
QTc interval delta vs. HCQ Cumulative Dose or Serum Level. The relationship to the QTc delta from the same day as the cumulative HCQ dose is shown Figure 4A. The relationship between QTc delta and HCQ serum levels is shown in Figure 4B. Each figure includes the red, linear regression line with equation.

79.2% had ≥ 1ECG while also receiving AZM. Again, there was no correlation between HCQ cumulative dose and QTc delta (r = 0.147). Four patients (16.7%) had a maximum delta QTc > 100 ms. All were continued on HCQ. Despite the increasing HCQ cumulative dose, the QTc delta went down in each patient and, in some cases, went below zero. No patient experienced torsades de pointes. Three patients (12.5%) were administered amiodarone while receiving HCQ and AZM. None had significant change in their QTc delta while exposed to amiodarone and HCQ & AZM.

Serum HCQ levels were measured on the same day as 48 of the ECGs analyzed. The HCQ level and QTc delta were not correlated (r = −0.078 & r_s_ = −0.031) (Figure 4B). Four of the 48 QTc deltas were > 60 ms and the four were associated with HCQ levels = 36, 84, 227 and 339 ng/ml.

## Discussion

### Outcomes

In this cohort of 255-IMV requiring Covid patients, the mortality rate was 78.8%. This rate is similar to published data on critically ill Covid patients from February through June 2020 (14–17) but lower than that of other studies. However, many studies on critically ill, IMV Covid patients reported only short-term outcomes and often, at the time of data censoring, these studies had significant percentages of patients still on the ventilator or hospitalized (18–23). In our cohort, at discharge or Day 90, only 16.7% of the Alive patients (3.5% of the Cohort) was discharged to home without any cognitive or motor deficits and off oxygen therapy.

### Demographics, Presenting Illness & Comorbidities

As noted in all prior reports on Covid, age was inversely correlated with survival; patients over the age 60 years had a 5-times greater chance of dying. As seen previously at this hospital (24), black was the most common race, unlike studies in nearby medical centers (25)(19,23,26). Race was not associated with outcome. Although only 16.7% of the Cohort had one or more of these conditions, active cancer, dementia, history of CVA and COPD, each expired. patients with history of renal transplantation had a high mortality rate (83.3%).

HTN, atrial fibrillation, coronary artery disease, and ESRD on renal replacement therapy have been associated with poor outcomes in cohorts of hospitalized or ICU Covid patients (25). However, these pre-existing conditions were not associated with outcome in this cohort. HLD was the only comorbidity associated with outcome. HLD patients had >3 times increased chance of death.

As previously observed, time to intubation was not associated with outcome(21,26). However, patients, who sought medical attention prior to being admitted, were more likely to die than those who were admitted on initial presentation. This group also had a longer duration of symptoms prior to admission, but their NEWS2 scores were equivalent. This observation suggests that hospitalization changes the natural history of the disease.

### DM, Hyperglycemia & other Laboratory Parameters

Overall, the cohort had high rate of DM (59.2%). This rate is similar to some reported rates, (19,23,27) but higher than many. This difference can be explained partially because most prior observational studies, the providers did not actively attempt to diagnose DM in patients with hyperglycemia. an On admission, 45.5% had known DM; another 13.7% were diagnosed with DM, based on A1C value, during the admission. Not all patients without a known DM had an A1C measured unfortunately. Slightly less than 50% of patients no history of DM had an A1c measured. Of these, ∼90% had DM or PreDM. DM status was not associated with outcome.

The DM status remains unknown 29.0%, the vast majority (91.2%) of whom were hyperglycemic (BG > 140 mg/dl) in the absence of corticosteroid therapy. Since critical illness is often associated with “stress hyperglycemia”, we did not attempt to diagnose DM in this group of patients. However, based on the A1C results, a majority of these patients most likely had undiagnosed DM or PreDM. In fact, only 3.9% of the Cohort had BGs < 140 mg/dL throughout their admission.

Most initial laboratory values were not associated with outcome. However, the patterns of ferritin, D-dimer and LDH over time were strongly associated with outcome. The slope of each in the Expired group was several fold greater than that of the Alive group. Other researchers are starting to consider the trends of these markers for prognostic purposes(28). These results should be confirmed prospectively and could lead to better prognostication of Covid patients. AKI, as reported in many reports, was associated with an increased risk of death, and occurred 1.5 times more often in the Expired group.

### Obesity

Obesity, as observed in many studies, was common (50.4%) in this Cohort. Only 18.1% had a normal BMI, while 31.9% was overweight. This cohort’s mean BMI = 31.4. While similar to other reports on critically-ill Covid patients, this mean BMI is substantially higher than that seen in intravenous antibiotic trials.

Confirming prior results on hospitalized Covid patients, obesity was more common and more severe in younger patients in this Cohort (11). The mean BMI of patients < 60 years-old was 4.6 greater than that patients over ≥ 60 years-old. Accordingly, patients <60 years-old had a higher percentage of patients with severe obesity (BMI > 40).

BMI controls for height. However, the absolute weight range of the Cohort was extremely wide. The greatest weight was nearly 7-times the lowest. Further, the weights of 16.1% were ≥ 2 times the weights of 7.8% of the Cohort. These vast differences in weights would be expected to have a large impact on therapeutics, since the majority of medications was not weight-adjusted dosed.

### Therapeutics

As described above, all therapeutics were evaluated for correlation with outcome. However, few drug classes had any signal of association with outcome. Remdesivir was not available at this hospital during this period.

The Cohort was diagnosed and treated before publication of the Recovery Trial’s data on dexamethasone therapy (9). Using Recovery Trial’s dose of dexamethasone as cut-off dose, we analyzed the effect of corticosteroid treatment. and found no clear effect. Steroid use was more common in the Expired than the Alive group (50.2% vs. 37.0%).

Convalescent plasma (CP) has been studied in hospitalized Covid patients. Observation and RCT data on CP are mixed (29,30). In some studies, use of CP with high titers of antibodies against SARS-CoV-2 early in disease has been shown to improve survival. CP became available in mid-April through the Mayo Clinic program and was given to 19.6% of the Cohort. the antibody titers of each CP units are not known. Most patients received CP late in their disease course. The median day of CP administration = 10.8 days. While the survival rate for patients not receiving CP was much lower than those who did, the immortal time bias of late administration was evident. At 10 day, the survival rate for those not receiving CP increased to 37.8%, a rate higher than the survival rate of CP-receiving patients.

Tocilizumab (TOZ) was given to 35.7% of the Cohort. The data from observational studies and RCT trial on TOZ’s effect on Covid are also mixed (21–28). The overall TOZ-group survival rate = 35.2%. However, for similar aged patients, who did not receive TOZ, the survival rate after Day 5 of hospitalization or the median day of TOZ administration = 33.7%. Multivariate logistic regression analysis revealed TOZ therapy resulted in a 2.8 increase in survival. The effect of late administration to younger patients interferes with the interpretation of this apparent benefit.

Nearly 90% of the cohort received ≥400 mg HCQ. When prescribed, HCQ was begun early in the hospitalization. 86.1% received the first dose of HCQ within 24 hours and 94.2% within 48 hours of arrival. Several patients received their first dose of HCQ before they were officially admitted. The average age of HCQ patients was not different from those who did not receive the drug. Consequently, HCQ therapy data are not affected by immortal time and selection biases to the same degree as the CP and TOZ therapy data are. The dose and duration of HCQ therapy were determined by the Infectious Diseases (ID) consultant following a given patient. In March-April 2020, Covid patients were assigned to the one of the several ID groups based on a predetermined call schedule. Variations in dose and duration of HCQ were significant and dependent on the ID groups following the patients. The most commonly prescribed HCQ regimen (2,400 mg over 5 days) was given to 38.4% of the Cohort (31). The majority (77.2%) of the cohort received ≤3,000 mg total HCQ. Only 22.8% received >3,000 mg HCQ. Weight-adjusted dosing was rarely used.

AZM was co-administered to 62.5% of patients receiving HCQ. The decisions to co-administer AZM were made primarily by the ID consultants. The use of AZM with HCQ was more common in the first half of this 6-week period. During mid and late April, AZM was administered less frequently with HCQ. This decrease occurred after reports that HCQ/AZM caused QTc prolongation were published.

No randomized, clinical trial (RCT) has examined the effect of HCQ/AZM therapy in critically ill patients. Further, no other study, observational or RCT, has examined the relationship of a patient’s cumulative doses of HCQ and AZM to outcome. In this Cohort, the treatment with HCQ cumulative dose > 3 gm with AZM > 1 gm was associated with a much higher survival rate than patients who did not receive this combination. 37 patients received this “Treatment” and 218 patients received ≤ 3 g HCQ and/or ≤ 1 g AZM. The survival rate for the “Treatment” group = 48.6% and that for all others = 16.5%. Several analyses were performed to better understand what factors led to a relative increase in survival rate = 198%. After controlling for other covariates, including CP, steroids, and TOZ, the effect of this “Treatment” on survival rate was still great.

A Rubin causal analysis established that the “Treatment” (> 3 g HCQ/> 1 g AZM) increases the chances of survival by an absolute 22.5%. In other words, > 3 g HCQ/> 1 g AZM therapy increases the survival rate from 16.5% to 39.0% [p = 0.003]. This increase represents a 136% relative increase in survival rate.

The Rubin model established higher doses of HCQ and AZM greatly improved survival in IMV-requiring Covid patients. If correct, then weight-adjusted HCQ cumulative dose would, at some level, have a greater impact. In accordance with this hypothesis, an HCQ cumulative weight-adjusted dose > 82.2 mg/kg and > 1 g AZM produced an ATE = 46.1% [p=00.12].The fact that weight-adjusted cumulative dose has an even greater effect on survival than cumulative HCQ dose is strong confirmation of the causal relationship between this treatment and improvement in survival rate. Finally, the OR ratio for weight-based HCQ dosing was over twice that of a cumulative HCQ dose. Together, these analyses are consistent with each other and establish that HCQ/AZM, when given at relatively increased doses, has a large impact on survival in critically ill Covid patients who require IMV.

HCQ tissue levels are not only affected by cumulative dose, but also by the patient’s weight (32). As obesity is a risk factor for severe Covid, the mean BMI of critically ill Covid cohorts is typically > 30. HCQ regimens used in Covid clinical trials have not been adjusted for increased BMI or weight. Younger, hospitalized Covid patients have greater BMIs than older Covid patients, who often have normal or low BMI. BMI normalizes for height. Few Covid studies report on the absolute weight of patients. In the Cohort, the maximum BMI was 3.8 times the minimum, while the maximal weight was 6.8 times the lowest weight. The ranges of both BMI and weight in Covid cohorts are greater than that seen in most antibiotic clinical trials. For instance, the Cohort’s average weight (89.7 kg) is 21 kg greater than that of the Merino trial (68.3 kg) (33). Further, the range of weights is greater in Covid patients. The Cohort’s S.D. for weight (25.2) is far greater than S.D. of the Merino trial subjects (18.7). In short, severely ill Covid patients have much greater weight averages and weight ranges than are typically seen in antibiotic clinical trials. Still, no Covid study has examined the effects of these differences in weight on response to HCQ therapy or HCQ adverse effects.

Several reasons explain the inconsistent results from clinical trials. HCQ’s half-life (*t*_1/2_) is >40 days. This *t*_1/2_ is much greater than most, if not all, commonly used antibacterial and antiviral agents. HCQ’s proposed mechanism of action against SARS-CoV-2 is indirect. HCQ is not thought to interfere with viral functions directly, but, by affecting the pH of lysosomes, HCQ reduces the chance a cell can be infected. In the collagen-vascular disease literature, it is well established that HCQ’s effect on symptoms does not reach maximum until many weeks to months of daily HCQ administration. The maximal effect is associated with “build up” of HCQ in tissues. Given these facts, it is expected that HCQ’s effect on Covid would be dependent upon accumulation of HCQ in lungs and respiratory tract tissues. Given the wide range of weights and BMIs in Covid patients, the minimally effective cumulative dose would most probably vary significantly by weight/BMI. Therefore, effective HCQ cumulative dose of a lean population would be lower than that of a population with greater BMIs/weights. In other words, trials with leaner cohorts might have better results.

Lead-in or dose-escalation studies in rheumatoid arthritis (RA) and systemic lupus erythematosus (SLE) patients have established that HCQ, given at dose higher and for much longer than those used to treat Covid, are safe and well tolerated. In the studies by Costedoat-Chalumeau *et al*. and Furst *et al*., SLE and RA patients were given daily doses of HCQ up to 800 mg or 1,200 mg for 6 weeks or more (34,35). The adverse effect profiles for the highest doses were not different from those of lower doses. In these studies, there was no suggestion of cardiotoxicity from higher doses of HCQ.

Our findings, while unique, are supported by findings of several observational or single arm studies (36–42). Lagier *et al*. reported that HCQ/AZM therapy was associated with improved outcomes in a large, observational study. Lauriola *et al*. similarly showed that HCQ with AZM was associated with a large increase in survival in hospitalized Covid patients compared with HCQ therapy alone. Our analyses suggest that AZM is not simply a marker of higher dose HCQ but contributes to improvement on survival.

### Effect of HCQ +/-AZM on the QT Interval

Since the survival benefit was great, the effect of HCQ on the QT interval on this Cohort is not clinically important. However, for other populations, this issue is relevant. HCQ is the drug of choice for SLE and often used to treat RA patients. These patients take HCQ daily, usually for decades. Analysis of HCQ’s effect on QTc in SLE pt was negative. However, HCQ therapy of Covid has been linked to prolongation of the QT interval. The reports, which showed an increase in the QTc with HCQ therapy, were uncontrolled. Two studies analyzed the greatest increase in QTc while a patient was on HCQ therapy (43–45). Typically, the association of a specific drug with QTc prolongation is evaluated using pharmacodynamic-pharmacokinetic modeling. None of the reports, which claim HCQ causes QTc prolongation, used PD/PK modeling (46). One study reported the effects of HCQ therapy over a the 5-day course of therapy (45). HCQ blood levels were not assessed. The total dose of HCQ given was only 2,400 mg. We compared the QTc delta to cumulative doses much greater than 2,400 mg and to serum HCQ levels.

The Cohort’s data establish that, even with AZM co-administration, HCQ therapy and QTc interval changes are not correlated. Of note, the peak QTc increase in the Cohort patients often occurred early in the course of HCQ therapy and before the minimum QTc delta occurred. As the patient improved clinically, the patient’s QTc delta decreased. Similar to cumulative HCQ dosages, serum HCQ levels are not associated with QTc changes. Together, these data suggest that QTc prolongation seen in Covid patients is secondary to underlying illness and not HCQ therapy.

On April 24, 2020, the FDA issued a warning about the possible effects of low HCQ on QTc interval (47). Since 2010, the FDA has approved over 150 clinical trials, which include HCQ treatment. The FDA did and does not require monitoring for cardiotoxicity. In each of these trials, the total HCQ dose and expected tissue levels are markedly higher than used or seen in Covid patients. This discrepancy lacks logic or explanation.

In studies on drug-induced QTc prolongation, the QTc improves when the drug is discontinued. However, in these reports, as in this Cohort, the maximal QTc change often occurred days before HCQ was discontinued. The trials, which concluded HCQ caused QTc prolongation, did not show that a dose-based effect. Given the weight range of hospitalized Covid patients, it is difficult to understand how weight-adjusted dosing was not considered in these reports. At a minimum, the studies on HCQ’s effect on QTc interval in Covid patients need to be redone and the extant data re-analyzed.

## Conclusions

This study extends the understanding of outcomes in Covid patients, who require IMV. Only 3.5% of the Cohort “walked out of the hospital”. While several factors have been associated with outcome in hospitalized Covid patients, few are associated with outcome in Covid patients requiring IMV.

Most Covid studies have not considered days of therapy, cumulative dose, or weight-adjusted dosing. We found that when the cumulative doses of two drugs, HCQ and AZM, were above a certain level, patients had a survival rate 2.9 times the other patients. By using causal analysis and considering of weight-adjusted cumulative dose, we prove the combined therapy, >3 g HCQ and > 1g AZM greatly increases survival in Covid patients on IMV and that HCQ cumulative dose > 80 mg/kg works substantially better. These data do not yet apply to hospitalized patients not on IMV. Since those with higher doses of HCQ had higher doses of AZM, we cannot solely attribute the causal effect to HCQ/AZM combination therapy. However, it is likely AZM does contribute significantly to this increase in survival rate. Since higher dose HCQ/AZM therapy improves survival by nearly 200% in this population, the safety data are moot. However, given the data presented here, the studies reporting HCQ’s effect of QTc intervals need to be re-evaluated.

## Data Availability

Data were extracted and combined into a database. Each patient's data were combined into a dataset containing relevant time, date, result, and dosing information. In addition, each chart was reviewed to extract past medical history, prior medical attention, and outpatient medication use. These are available upon request in large format time series datasets in CSV filetype.

